# Role of Covid vaccine in determining ICU admission and death due to Covid-19 in Tamil Nadu

**DOI:** 10.1101/2022.02.07.22270596

**Authors:** T S Selvavinayagam, K Parthipan, Sudharshini Subramaniam, A Somasundaram, P Sampath, Vinay Kumar

## Abstract

COVID-19 pandemic threatened the world in terms of its rapid spread, strain on health infrastructure and many people lost their lives due to COVID. Mass Vaccination of public against Covid-19 were done with the notion that it protects against the severe form of the disease and death due to Covid-19. Covid vaccination was rolled out in Tamil Nadu from 16^th^ January 2021 in a phased manner. This study was done using secondary data to assess the role of COVID vaccination in preventing ICU admission and death due to Covid-19 in Tamil Nadu for the period of August – December 2021. Unvaccinated individuals contributed to a higher proportion of hospitalization (60.9%) and ICU admission (65.5%) among Covid-19 infected during this period. Similarly, among patients who died due to Covid-19, 75.5% were unvaccinated. Odds of ICU admission and death among unvaccinated was 2.01 and 3.19 - **times** higher compared to fully vaccinated individuals infected with Covid-19. Unvaccinated Covid-19 patients had **2.73- and 1.46-** times increased odds of dying and ICU admission respectively, compared to partially vaccinated. Population Attributable Risk showed that receiving at least one dose of vaccine could have reduced the mortality among Covid patients by 54% and ICU admission by 23.3%. This article emphasizes the need for vaccination against Covid-19 to reduce ICU admission and death among those infected with Covid-19.

## Introduction

Ever since the 1^st^ case of Covid-19 reported in China, the world has been facing the impact of Covid-19 with no countries being spared. World has witnessed the rise in Covid cases in different phases, which is usually referred to as *waves*. In the 1^st^ wave, which lasted for almost 9 months, public health measures were focussed on quarantine, isolation, social distancing, wearing of mask and containment. Efforts to reduce the morbidity and mortality due to the pandemic was aided by mass lockdown measures. Countries were naïve to this virus; thus, the prevention and management measures were all learnt through trial and error. Vaccines to protect against Covid-19 was a felt need across the globe, as the pandemic was not only affecting the health but the entire socio-political economy due to the lockdown. Clinical trials for different vaccines using different technologies were carried out in different parts of the world. Vaccines were rolled out for use towards the end of 2020. While the world was taking a breath of relief with the vaccines introduced, the 2^nd^ wave reportedly dominated by the delta variant of the Covid virus had already begun. Vaccines against Covid-19 seem to be protecting against the severe form of the disease and death due to Covid-19. ^1,2^

Tamil Nadu, the southern state of India, reported its first case of Covid-19 on 7^th^ March 2020. Ever since then, the number of cases continued to increase with the 1^st^ wave reaching its peak in the month of July,2020. Covid vaccination was rolled out in the state from 16^th^ January 2021 with Covaxin and Covishield introduced for Health care workers, followed by front line workers, >60 years, 45-60 years, 18-45 years in that order. It was from the month of March 2021, the state started witnessing the beginning of the second wave. As per the Indian Council of Medical Research(ICMR) guidelines, testing-tracking-tracing, isolation and home-based treatment of positive patients was the key measure to curb transmission of SARS-CoV-2.^3^ Unlike the 1^st^ wave, there were large number of cases infected in a short period of time which strained the health system. There was a high demand for admission to Intensive Care Unit (ICU) and oxygen during this wave. The fear of Covid-19 was more visible in the 2^nd^ wave than the 1^st^, which was further perpetuated by the high number of deaths being reported. This acted as a demand generator for Covid-19 vaccination in the later months. In Tamil Nadu, as on December 31, 2021, 59% of the eligible population have been fully vaccinated and 86% are partially vaccinated. There still exist a larger proportion of people who are yet to be fully vaccinated.

This article is an attempt to understand the real-world impact of vaccination in ICU admission and Case fatality among Covid infected people in Tamil Nadu.

## Methodology

The study was done using the secondary data obtained at the State Emergency Operations Control Room on hospitalization, ICU admission, death, and vaccination status. This data was collected from all government and private hospitals, medical colleges, covid care centres (CCC) of Tamil Nadu from the month of August 2021 to December 2021. Data on home isolation for this period was obtained from the office of the Deputy Director of Health Services of the concerned Health Unit Districts and City Health Officer of Greater Chennai Corporation. This period was chosen, as 2^nd^ wave was declining and there was an increase in the uptake of vaccination among general population.

Any admissions in CCC during this period was done only for patients who were otherwise normal but did not have adequate facilities at home for isolation.^4^ Hence, for the purpose of this study, admissions in CCC and direct home isolation was considered as non-hospitalization. Any admissions reported from any hospital (government or private hospital) was considered as hospitalization. Those who required ICU care during the course of treatment were considered as ICU admission. All death cases who were RT-PCR positive for SARS CoV-2 were considered as COVID deaths.

## Results

During the period of August-December 2021, there was data on 2,15,501 people who were infected with Covid-19, of whom almost 57% were unvaccinated, 25% were partially vaccinated and 18% fully vaccinated. Among the infected, 37.1% required hospitalization, 5.25% required ICU care and the Case fatality rate was 1.2%.

Figure 1,2,3, shows the vaccination status of the various parameters namely, hospitalization, ICU admission and death. It is found that in all the outcomes studied, a larger proportion was contributed by the unvaccinated individuals, followed by partially vaccinated. (Figure 4)

**Figure 1.**
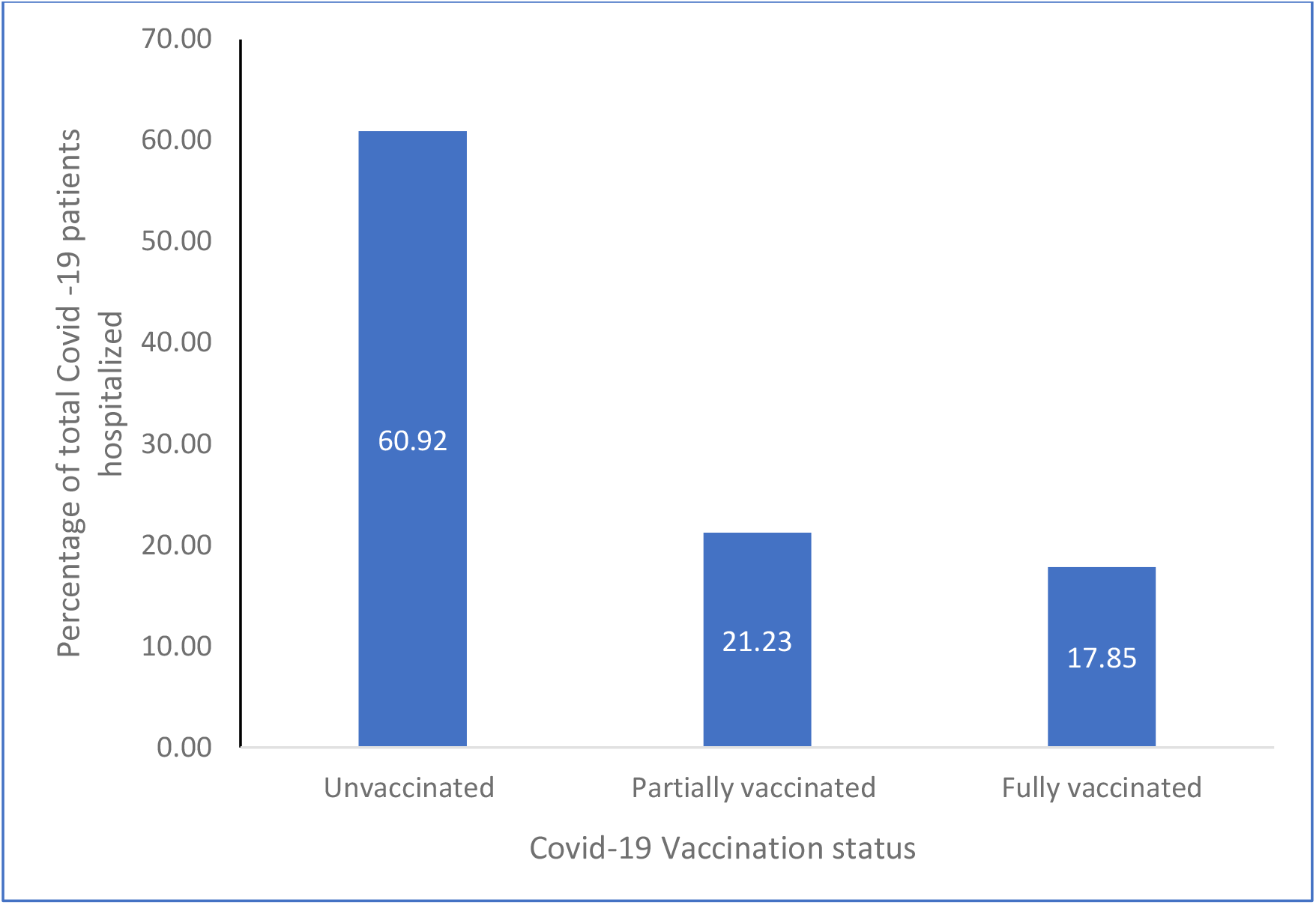
Covid-19 vaccination status among patients hospitalized due to Covid-19

**Figure 2.**
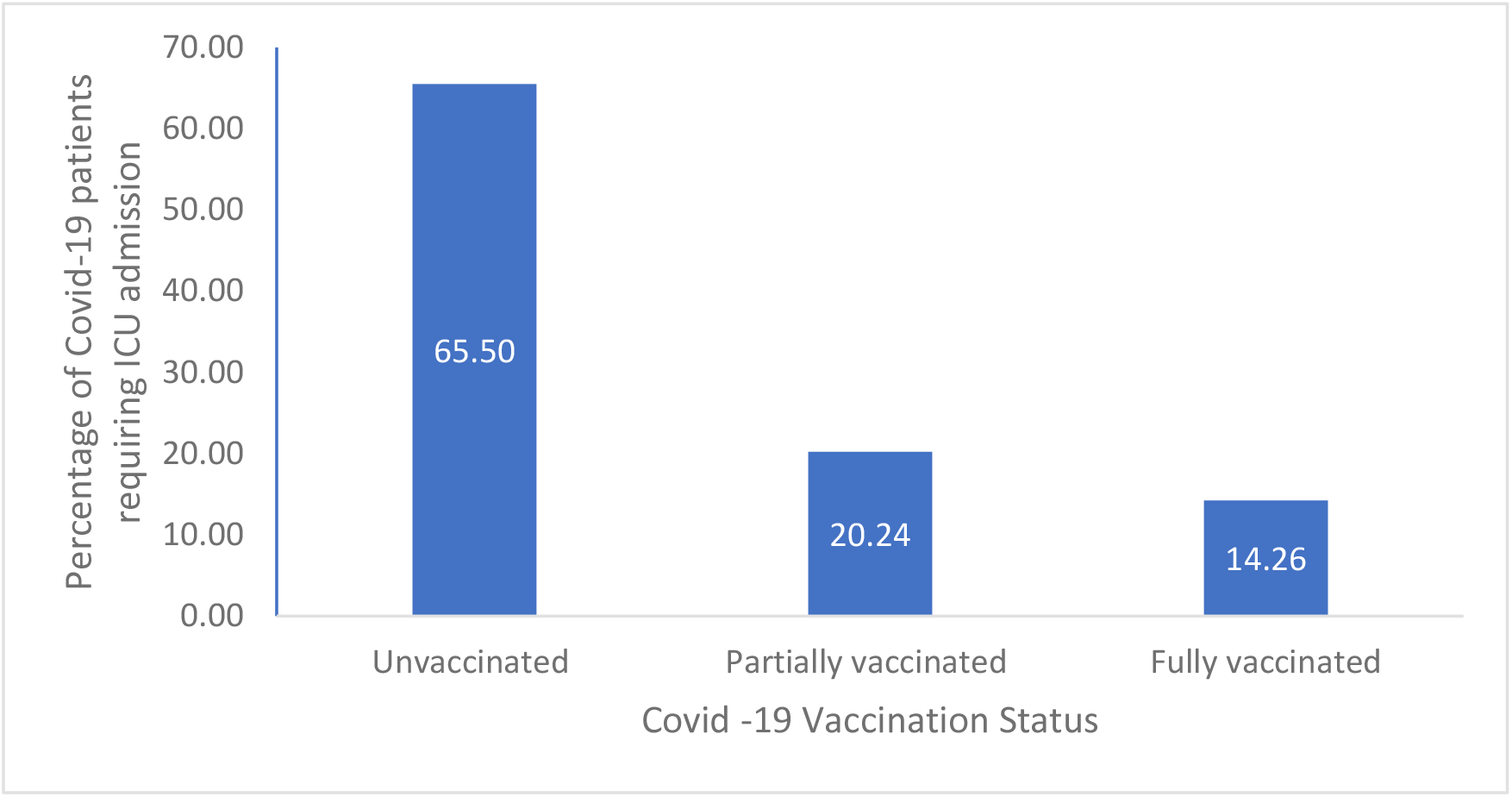
Covid-19 vaccination status among ICU admissions due to Covid-19

**Figure 3.**
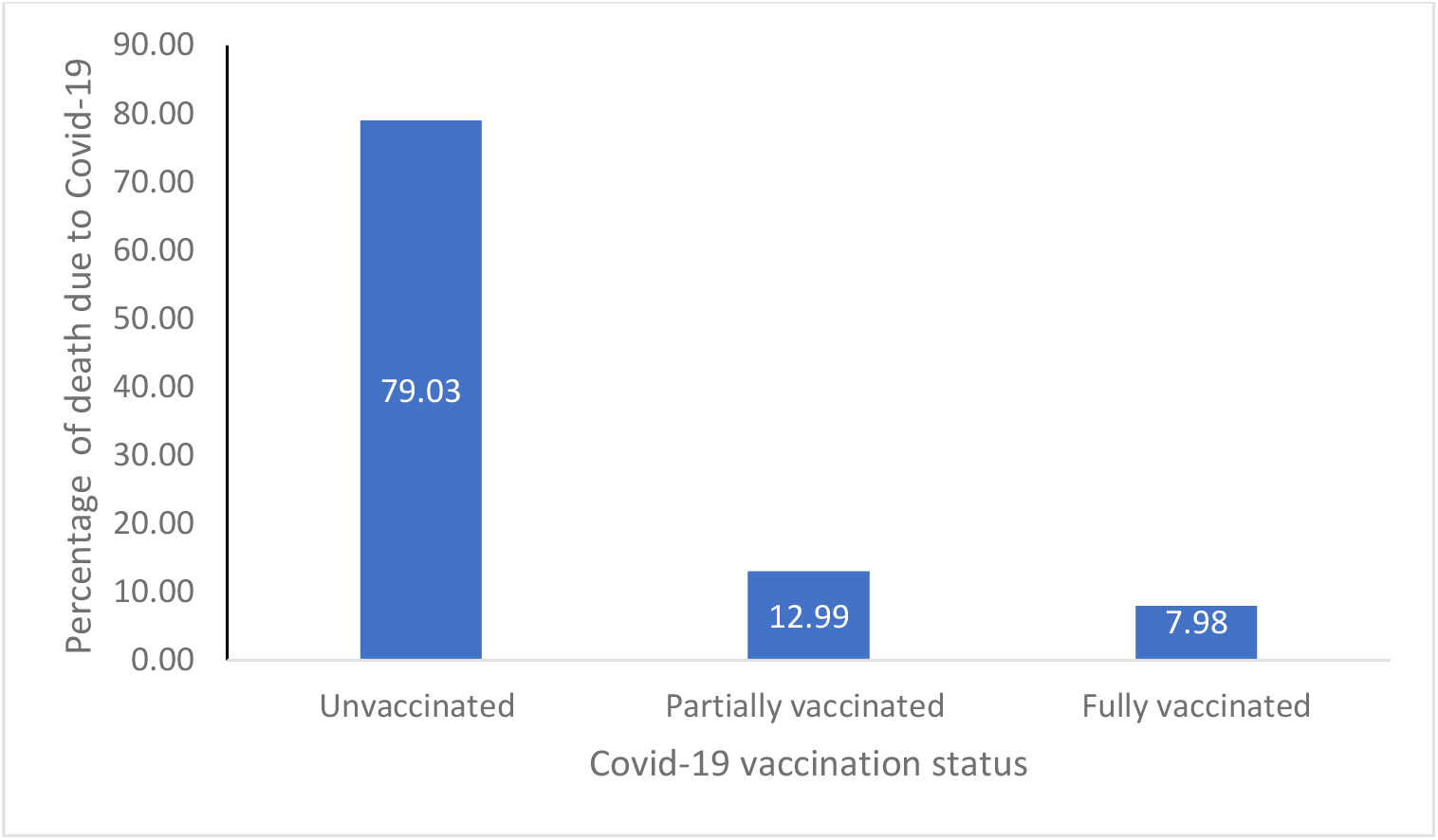
Covid-19 vaccination status among deaths due to Covid-19

**Figure 4.**
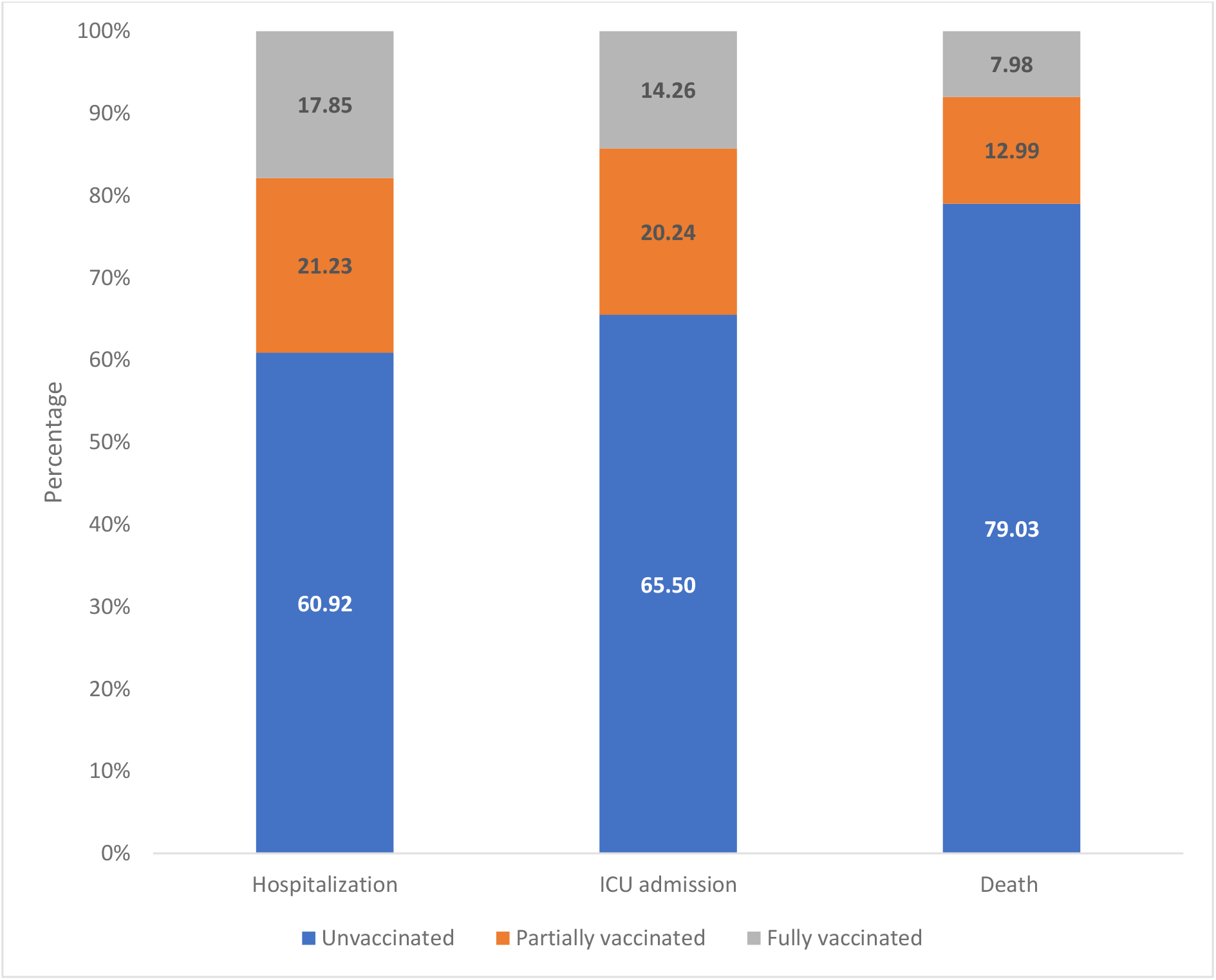
Covid vaccination status across different outcomes among Covid-19 patients

Figure 5 gives the stratified analysis of ICU admission rate and case fatality rate based on vaccination status among Covid-19 patients. Unvaccinated individuals had a higher ICU admission rate and case fatality rate compared to fully /partially vaccinated.

**Figure 5.**
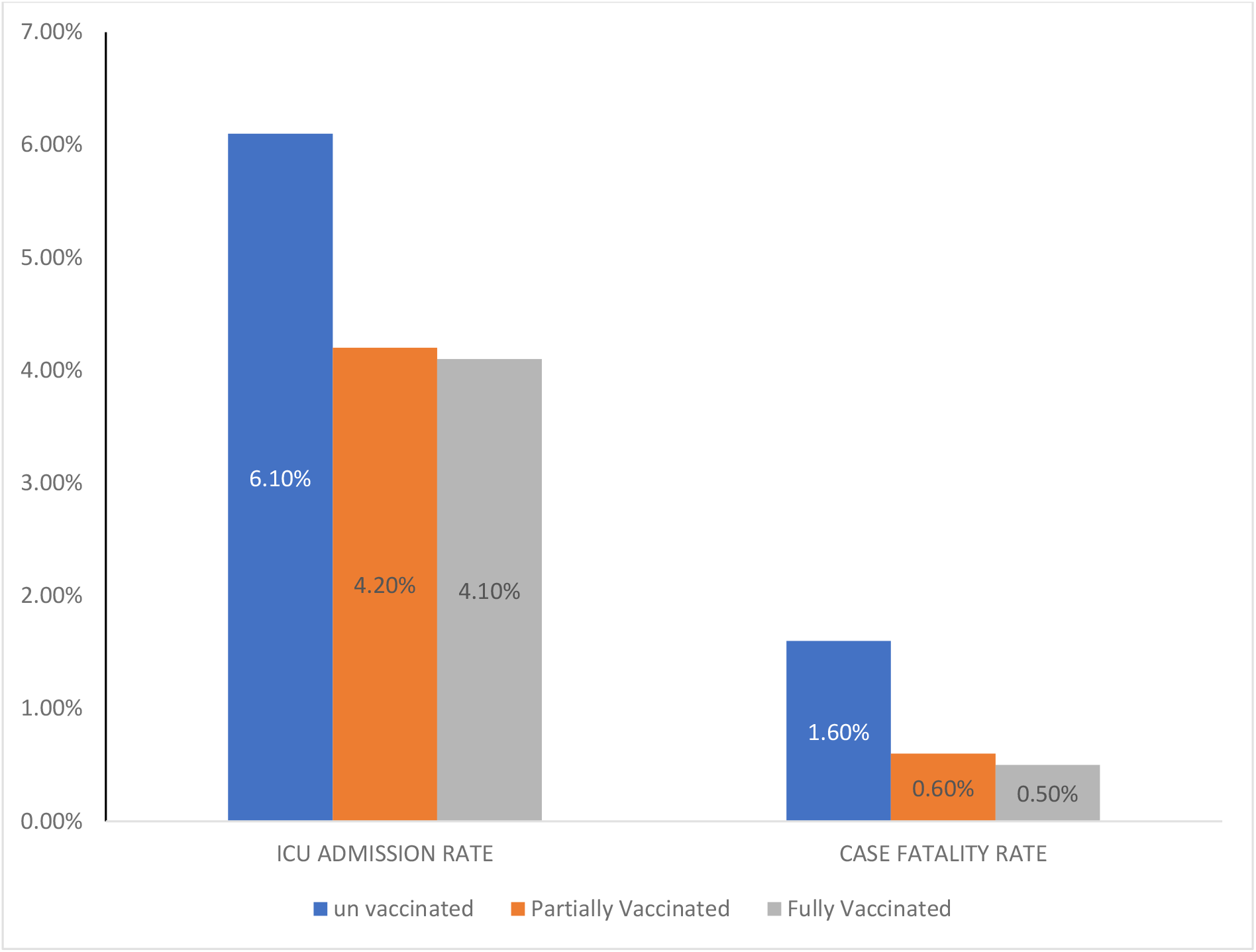
ICU admission rate and Case Fatality Rate among Covid-19 patients based on vaccination status.

Table 1 and 2 gives the Odds Ratio, Attributable Risk and Population Attributable Risk of case fatality and ICU admission among Covid-19 patients. Odds of ICU admission among unvaccinated was 2.01**times** higher compared to fully vaccinated individuals which was statistically significant. However, the odds of ICU admission among partially vaccinated compared to fully vaccinated was 1.01, which was not statistically significant.

**Table 1.** Case fatality among Covid patients - Comparison of unvaccinated and partially vaccinated with fully vaccinated.

| Covid vaccination status | Case fatality among Covid patients |  |  |  |
| --- | --- | --- | --- | --- |
|  | OR (95% CI) | P value | AR (95% CI) | PAR |
| Unvaccinated vs Fully Vaccinated | 3.19(2.76-3.68) | <0.0001 | 68.35% (60.36-76.33) | 62.08% |
| Partially vaccinated vs Fully Vaccinated | 1.16(0.98-1.39) | 0.077 | 14.37% (-1.6-30.3) | 8.9% |

OR- Odds Ratio; CI – Confidence interval, AR – Attributable risk, PAR – population Attributable risk

**Table 2.** ICU admission among Covid patients - Comparison of unvaccinated and partially vaccinated with fully vaccinated.

| Covid vaccination status | ICU admission among Covid patients |  |  |  |
| --- | --- | --- | --- | --- |
|  | OR (95% CI) | P value | AR (95% CI) | PAR |
| **Unvaccinated vs Fully Vaccinated** | 2.01(1.19-2.12) | <0.0001 | 50.4% (46.7-54.1) | 41.4% |
| **Partially vaccinated vs Fully Vaccinated** | 1.01(0.95-1.08) | 0.577 | 28.6% (23.3-33.9) | 16.7% |

OR- Odds Ratio; CI – Confidence interval, AR – Attributable risk, PAR – population Attributable risk

Similarly, unvaccinated individuals had **3.19-** times increased odds of dying compared to fully vaccinated individuals which was statistically significant. However, the odds of dying among partially vaccinated was not statistically significant compared to fully vaccinated.

Compared to fully vaccinated, attributable risk of death and ICU admission due to non-vaccination among Covid patients is 68.35% and 50.4% respectively and is statistically significant, which means that 68.35% of deaths and 50.4% of ICU admissions among unvaccinated could have been prevented if they had been fully vaccinated.

From table 3, it is found that unvaccinated Covid-19 patients had **2.73- and 1.46-** times increased odds of dying and ICU admission respectively, compared to partially vaccinated which was statistically significant. Attributable risk of death and ICU admission due to non-vaccination compared to partial vaccination was 63% and 30.5% respectively and is statistically significant, which means that 63% of deaths and 30.5% of ICU admission among unvaccinated could have been prevented if they had received at least one dose of vaccine.

**Table 3.**
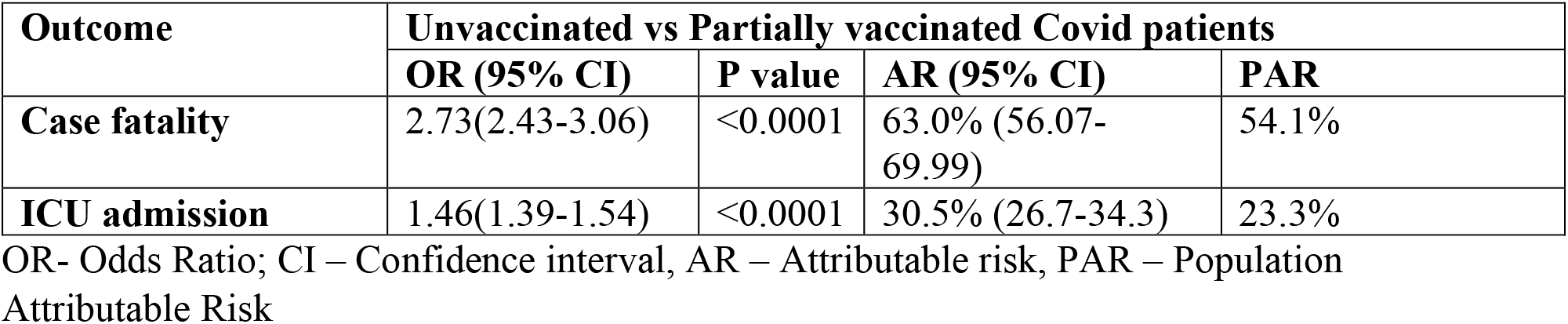
Case fatality and ICU admission - comparison between unvaccinated and partially vaccinated Covid patients.

| Outcome | Unvaccinated vs Partially vaccinated Covid patients |  |  |  |
| --- | --- | --- | --- | --- |
|  | OR (95% CI) | P value | AR (95% CI) | PAR |
| **Case fatality** | 2.73(2.43-3.06) | <0.0001 | 63.0% (56.07-69.99) | 54.1% |
| **ICU admission** | 1.46(1.39-1.54) | <0.0001 | 30.5% (26.7-34.3) | 23.3% |

OR- Odds Ratio; CI – Confidence interval, AR – Attributable risk, PAR – Population Attributable Risk

## Discussion

It is evident from the state-wide data that vaccination against Covid-19 is protecting people from severe form of disease which is measured using surrogate indicators like Hospitalization, ICU admission and Case Fatality. Among hospitalized patients, 17.9% were fully vaccinated, 21.2% partially vaccinated and 60.9% unvaccinated. The protective effect of vaccination was more obvious as the severity of the disease increased, which is evident by the fact that among people requiring ICU admissions due to Covid-19, 65.5% were unvaccinated, 20.2% were partially vaccinated and only14.3% were fully vaccinated. Such protective effect of the vaccine was observed maximum against death. Among those who have died due to Covid-19, majority (79%) were unvaccinated, 13% partially vaccinated and only 8% were fully vaccinated.

Population Attributable Risk shows that receiving at least one dose of vaccine could have reduced the mortality among Covid patients by 54% and ICU admission by 23.3%. PAR also shows that receiving two doses of Covid vaccine could have reduced the mortality among Covid patients by 62.1% and ICU admission by 41.4%. This finding goes well with the model predicted by Moghadas et al using the United States data, which predicted 69.3% protection from death following vaccination. ^5^

This article has focussed only on the vaccination status, while other determinants of severity of disease and mortality like age, comorbidity, time interval between infection and admission, treatment received, etc. have not been accounted or adjusted in this analysis. We would also like to highlight the possibility of bias due to differential health seeking behaviour, as people who had already received vaccine are more health conscious and therefore more likely to seek care at the earliest.

Since the vaccination was introduced in phased manner targeting the vulnerable population earlier, there is a chance that vaccinated individuals are at more risk of infection and disease, which could also be a possible explanation for ICU admission and death among vaccinated. It should also be noted that this study has not considered the time interval between vaccination, occurrence of disease and outcome. Seroconversion following vaccination and the waning effect of vaccine induced immunity also plays a vital role in determining the protection against the infection and severity of disease. These factors have not been assessed in this study. A follow up Cohort study among vaccinated individuals is required to understand if the benefits of vaccine continue to give protection over longer period, need for booster doses of vaccine and effect of vaccine on changing strains of Covid-19.

## Conclusion

This article gives information on real world effect of vaccine in reducing ICU admission and Case Fatality in Tamil Nadu and emphasizes the need for vaccination against Covid-19 to reduce ICU admission and death among those infected with Covid-19. While, having at least a single dose of vaccine has an impact on reducing ICU admission and death rate compared to no vaccination, it should be also emphasized that getting two doses of vaccination had a higher protective effect.

## Data Availability

All data produced in the present study are available upon reasonable request to the authors

